# Clinical performance and safety of Cerviron^®^ vaginal ovules in the local treatment of non-specific vaginitis: a national, multicentric clinical investigation

**DOI:** 10.1101/2023.05.09.23289715

**Authors:** Daniela Oana Toader, Raluca Alexandra Olaru, Dominic-Gabriel Iliescu, Ramona Petrita, Florentina Liliana Calancea, Izabella Petre

## Abstract

**Purpose:** Non-specific vaginitis is a distinct clinical entity with particular microscopic and immunologic features. Currently, there is no standard of care for women with non-specific vaginitis. The aim of our study was to assess the change in vaginal symptoms score after a 3-months treatment with Cerviron^®^ medical device in participants with abnormal vaginal discharge and with specific signs and symptoms. As secondary objectives, the study analyzed other clinical and microscopic features, such as vaginal discharge aspect, change in vaginal pH, change in vaginal microbiome, and vaginal inflammation.

**Methods:** The study population included 47 participants suffering from symptomatic vulvovaginitis, distinct from candidiasis, trichomoniasis or bacterial vaginosis. The study design included 2 research sites from Romania. The treatment protocol consisted of 1 ovule/day inserted intravaginally, during 15 consecutive days. The total study duration was 3 months.

**Findings:** Cerviron^®^ had a positive impact on the vaginal symptoms score for 72.34% of the study participants. Topical administration of Cerviron^®^ balanced vaginal pH values and significantly reduced signs of inflammation between study visits.

**Implications:** Cerviron^®^ shows curative effects that supports its use as a stand-alone treatment in women with non-specific vaginitis. ClinicalTrials.gov identifier: NCT04735705.

A second clinical investigation is ongoing to evaluate its clinical efficacy in postoperative care of cervical and vaginal wounds, traumatic or secondary to surgical interventions.

## 1. Introduction

### 1.1. Distinction between specific and non-specific vaginitis

Vaginitis is a complex gynecological condition, characterized by vaginal symptoms such as abnormal vaginal discharge, malodor, irritation, dysuria, dyspareunia and vaginal inflammation [1]. Vaginitis is encountered at all ages, from children and adolescents to postmenopausal patients suffering from vaginal atrophy [2,3]. The vaginal microbiome is a constantly changing ecosystem consisting of *Lactobacillus* and aerobic bacteria [4]. The imbalance of the vaginal species (also called a dysbiosis) triggers vaginal infections with abnormal vaginal discharge as the most frequent reported symptom. There should be a clear dissociation between specific vaginitis and non-specific vaginitis. Specific vaginitis is caused by aerobic microorganisms such as *Gardnerella vaginalis*, responsible for the majority of cases of bacterial vaginitis identified in clinical practice, with the second place occupied by *Candida species* causing vulvovaginal candidiasis and third, *Trichomonas vaginalis* causing trichomoniasis [5].

Non-specific vaginitis, on the other hand, is caused by an imbalance in the normal vaginal microbiome with the disruption of *Lactobacillus* and proliferation of gram-positive and gram-negative aerobic species such as *Corynebacterium, Staphylococcus epidermidis, Enterococcus faecalis, Escherichia coli (E. Coli), group A, B and D Streptococci* [6,7]. Non-specific vaginitis is also known as aerobic vaginitis due to the fact that the infection is caused by the proliferation of aerobic, endogenous bacteria. This type of vaginitis is usually misdiagnosed, although it has distinct clinical features such as vaginal inflammation, thinner vaginal mucosa and fewer superficial squamous epithelial cells with an increase in parabasal epithelial cells [8–10].

### 1.2. Adverse pregnancy outcomes related to aerobic vaginitis

Aerobic vaginitis poses a significant risk for negative pregnancy outcomes such as preterm rupture of membranes, preterm delivery, chorioamnionitis and funisitus of the fetus [8,11]. Misdiagnose (or confusion with bacterial vaginosis) and improper therapeutic strategy can translate into spontaneous miscarriage especially if the infection occurs in early pregnancy. Preterm birth was often associated with *E. Coli* and *Klebsiella* infections. *Staphylococcus aureus* presence in vaginal samples was associated with intrauterine growth restriction, while *group B Streptococci* increase the risk of maternal-fetal transmission of bacterial organisms. In neonates, the absorption of chorionic tissue by *Streptococci* led to pneumonia, sepsis and meningitis [12]. Donders et al. suggested a relationship between aerobic vaginitis and severe neurologic injuries such as neonatal cerebral palsy [13].

### 1.3. Other types of symptomatic vaginitis

Atrophic vaginitis, desquamative inflammatory vaginitis (erosive vaginitis), irritating/allergic vaginitis, and erosive vulvar lichen planus are types of non-infectious vaginitis. The etiology of non-infectious vaginitis can vary: the diminishing level of estradiol due to aging in the case of atrophic vaginitis, contact irritation or allergic reaction (caused by chemical products, latex or semen) or possible autoimmune mechanisms in the case of erosive vaginitis. Other miscellaneous causes can include trauma, foreign body or vesicovaginal fistula [15].

Non-infectious vaginitis is characterized by abnormal vaginal discharge and inflammatory features such as irritation, oedema and erythema [1,14,15]. The differential diagnosis implies exclusion of infectious microorganisms, determination of hormone levels and histology to identify the cause of vaginal inflammation.

### 1.4. Management of non-specific vaginitis

The purpose of treatment in both non-specific vaginitis and non-infectious vaginitis is to restore the vaginal microbiome and to reduce the signs and symptoms associated with inflammation. Several vaginal topical treatments were marketed until date to treat vaginal microbiome dysbiosis [16,17]. Among other potential interventions, vaginal microbiota transplantation shows the advantage of completely restoring the microbiome to a healthy state but there is still insufficient clinical evidence to translate this procedure into clinical practice [18]. Despite the therapeutic arsenal available in the market, the ideal duration of treatment or the standard of care have not been established yet. Moreover, treatment with antibiotics is inappropriate in the absence of microbiological tests and may have a negative effect on the vaginal ecosystem [19]. However, in selected cases antibiotics should be prescribed for short-term treatment, as longer treatments could induce antibiotic resistance [20]. Relapses after treatment are common and require maintenance therapy. Aerobic vaginitis is frequently associated with candidiasis or other mixed vaginal infections [21].

We designed the present multicentric clinical investigation (NCT04735705) to evaluate the clinical efficacy of Cerviron^®^ vaginal ovules as a stand-alone treatment in aerobic vaginitis.

## 2. Materials and Methods

### 2.1. Study Design

This study was designed as an open-label, pilot, multicentric, national clinical investigation. The study took place between 15 July 2021 (First Participant In) and 28 February 2022 (Last Participant Out) in 2 investigational sites located in Bucharest and Timisoara, Romania.

The purpose of this study (NCT04735705) was to investigate the therapeutic performance and tolerability of Cerviron^®^ in participants with aerobic vaginitis. The effects were assessed as changes between baseline and 3 consecutive study visits (planned at 30 days interval) in the following primary performance outcomes: clinical performance assessed by gynecological examination and the presence of vaginal symptoms assessed by the participant. The assessment of performance of the medical device included several other clinical outcomes, as follows: change in vaginal discharge aspect, change in vaginal pH values, change in vaginal microbiome, change in vaginal inflammation, and participant satisfaction at the end of the treatment. Safety was assessed through the rate of treatment-related adverse events in subjects participating in the clinical investigation.

### 2.2. The medical device

Cerviron^®^ (Figure 1) is a medical device manufactured by Perfect Care Manufacturing S.R.L. and authorized following the provisions of the Medical Device Directive 93/42/EEC.

**Figure 1.**
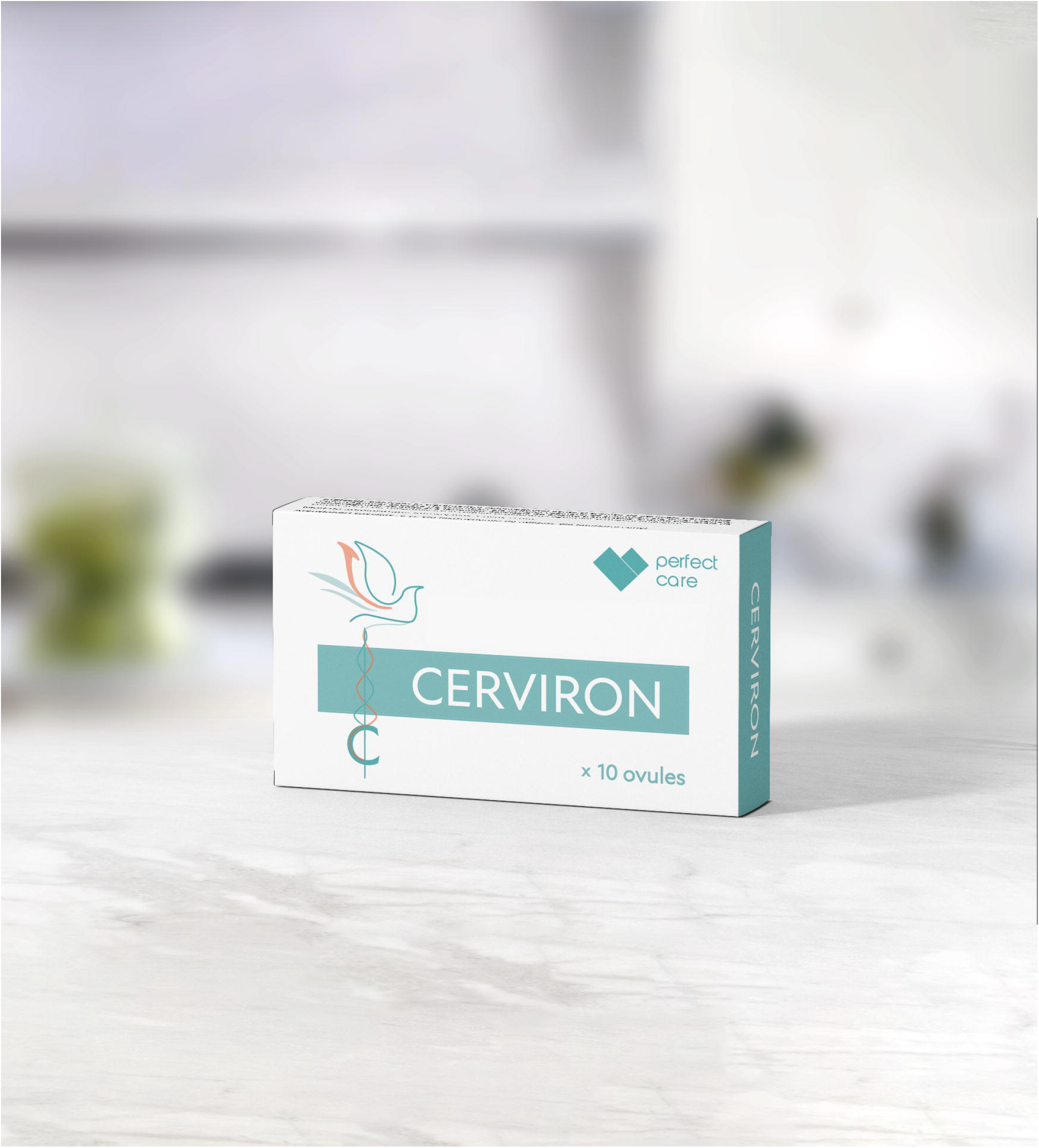
Cerviron secondary packaging.

Cerviron^®^ is marketed by Perfect Care Distribution in the following countries: Albania, Latvia, Lithuania, Estonia, Kosovo, Montenegro, Romania, Kuwait, and United Arab Emirates.

Cerviron^®^ has a complex composition consisting of three topical products – hexylresorcinol - 2mg, vegetable collagen – 15mg and bismuth subgallate - 100mg – and four phytotherapeutic extracts – *Calendula officinalis* – 10mg, *Hydrastis canadensis* - 10mg, *Thymus vulgaris* - 10mg and *Curcuma longa* - 10mg. Cerviron^®^ is formulated as homogeneous ovules, yellow to deep yellow, without undissolved particles or air bubbles. Each box contains 2 blisters with 5 ovules; the weight of an ovule is 2g.

Bismuth subgallate causes shrinkage of damaged tissue by stopping bleeding and promoting tissue healing. Being an insoluble salt and very little absorbed the bismuth subgallate forms a protective film that allows the other components to act locally. Collagen is structurally and functionally a key protein of the extracellular matrix which is also involved in forming the scars during the healing of conjunctive tissues, due to its chemotactic role. Many collagen bandages were developed to improve the repair of the wound, especially of non-infected, chronic, idle cutaneous ulcerations.The results of some studies indicate that methanol and ethanol extracts of *Calendula Officinalis* petals possessed good antimicrobial potential and antifungal activity [22,23]. The investigated plant extracts can be used for the preservation of processed foods, as well as pharmaceutical and natural therapies. In the composition of the medical device, *Calendula officinalis* is used to prevent contamination with exogenous bacteria during the handling of the medical device and to prevent microbiological contamination of the fat base with Gram-negative bacteria and fungi.

Cerviron^®^ is a medical device intended for use in the treatment of acute and chronic vulvovaginitis of atrophic, aerobic, traumatic, and infectious etiology, of cervical erosions of traumatic or infectious origin and of bleeding caused by them, in the postoperative recovery after gynecological surgeries, for restoration of pH and normal vaginal flora, as well as an adjuvant in treatment with antibiotics orantifungals.

Cerviron^®^ is indicated for the treatment of the following symptoms: leucorrhea, vaginal pruritus, burns in the vaginal area, rash, vaginal discharge with unpleasant odor, dysuria, dyspareunia and vaginal bleedings caused by a friable cervix.

Cerviron^®^ can be used, in association with a specific medicinal treatment, in the treatment of symptoms of vaginal infections with: *Gardnerella vaginalis, Trichomonas vaginalis, Candida albicans*, yeast cells and filaments.

Cerviron^®^ is contraindicated in case of hypersensitivity to any of the components of the product. Administration to pregnant women can only be done at the recommendation and under the supervision of a specialist doctor. No precautions need to be taken when used during breast-feeding. None of the components of the product affect the ability to drive or use machines.

Interactions with other products are not known, therefore it is recommended to consult a doctor before using Cerviron^®^ ovules together with other medical devices or medicines for vaginal use.

Cerviron^®^ should be stored at 2 - 25 ° C in the original package to protect from direct sunlight. The term of validity inscribed on the package refers to the product stored under the conditions provided by the manufacturer. The medical device should not be disposed of via wastewater or household waste.

The Instructions for use specify its field of use as adjuvant in the treatment of acute and chronic vulvovaginitis of mechanical etiology, caused by changes of vaginal pH and changes of the vaginal flora and of cervical lesions of mechanical origin.

The recommended use of Cerviron^®^ is one ovule per day, inserted on the first day after the menstruation and during 15 consecutive days. Its adjuvant role is consolidated during the administration for a period of 3 consecutive months.

This is the first clinical investigation in human subjects with this medical device.

### 2.3. Participant population

The target population for this clinical investigation was comprised of females aged 18-65 years old, suffering from symptomatic vulvovaginitis, of either non-infectious cause or aerobic vaginitis, with abnormal vaginal secretion and with characteristic signs and symptoms, such as pruritus, erythema, burning, pain, odor, dysuria and dyspareunia. Vulnerable subjects were not enrolled during this clinical investigation.

The National Medical Devices and Medicines Bioethics Committee (CNBMDM) approved the present clinical investigation with number 1DM from 01 April 2021 (protocol number CYRON/01/2021; NCT04735705). The clinical data were collected and reviewed in compliance with the ethical principles defined in the Declaration of Helsinki. Participants’ informed consent for enrolment in the study was signed before any study specific procedure took place.

### 2.4. Data collection

All data collected through the data management system, by electronic Case Report Forms, were checked for completeness and extreme values (outlier) presence. The data was collected using the electronic platform OpenClinica. OpenClinica is compliant with Food and Drugs Administration and European Medicines Agency regulations, including 21 Code of Federal Regulations Part 11, General Data Protection Regulation, The Health Insurance Portability and Accountability Act and International Council for Harmonization – Good Clinical Practice, as well as with data practices and security standards. Any anomalies found were forwarded by the study monitor to the Investigator. Such data queries were solved with priority, in no more than 3 days from the query’s opening. It was the statistician’s decision to use/accept the revised data for the subsequent statistical analyses. Data for the present clinical investigation is stored according to annex E of International Standard ISO 14155:2020 and can be made available upon request [24].

The collected data included population demography, medical history, collection of vaginal symptoms, collection of vaginal swabs, collection of vaginal smears (vaginal discharge), vaginal pH measurements, inflammatory and parabasal epithelial cells by microscopy and participant satisfaction after using the medical device (5-point Likert Scale).

The visit procedures included a microscopy evaluation of vaginal discharge, and wet mount (laboratory-based microscopy) at 30 days interval during 3 consecutive months. Eligibility check of each participant was performed by the treating physician after the collection of the vaginal swab at the screening visit and eliminating the suspicion of bacterial vaginosis, candidiasis or trichomoniasis. Results were provided through wet mount and Gram stain microscopy performed by local clinical laboratory from Institutul National pentru Sanatatea Mamei si Copilului “Alessandrescu-Rusescu” Bucharest (for clinical site 01) and private laboratory Bioclinica Timisoara (for clinical site 02). Vaginal pH measurements were performed using Vaginex^®^ pH strips, produced by Care Diagnostica Austria and distributed in Romania by Self Care Medical.

Vaginal tampons usage was prohibited during the clinical investigation because tampons can absorb some of the active substances and as such, they can prevent the ovule from exhibiting its full performance. Also, a list of restricted concomitant medications was defined in the clinical investigation plan for the purpose of distinguishing between the efficacy of the medical device in questions and other therapies. All concomitant medication and treatments were recorded in the appropriate study documents.

Adverse events and concomitant medications were collected at each visit. Each participant was followed for 3 months or approximately 90 days. The data was exported and analyzed using Analyse-it for Microsoft Excel (version 5.90, Leeds, UK).

### 2.5. Statistical analysis

Statistical analysis was performed using the R statistical software (version 4.1.1) with a few extra statistical packages added, all of which have been revised and updated to the latest version. The final analysis was completed after all subjects have finalized the study, all queries have been solved, and the database has been locked. The overall type I error was preserved at 5%; p-values < 0.05 were considered statistically significant, and 95% confidence intervals were calculated.

Statistical analysis was conducted on all subjects who have successfully completed the study without a protocol deviation that is regarded as impacting the assessment of the key variables (the Per Protocol Set population). To evaluate changes of proportions over time before and after the treatment, for categorical variables two-proportions Z-tests were performed. For primary outcome a test for equality of proportions with continuity correction, at 5% significance level was performed. McNemar’s Chi-squared test with continuity correction was performed to assess the status-quo of the vaginal discharge aspect score between the baseline and final visit.

## 3. Results

### 3.1. Characteristics of the Patient Cohort

Participants were aged between 22 and 64 years, of Caucasian ethnicity (typical of the Eastern Balkan region), with different baseline characteristics, as shown in Table 1. Out of 47 participants, 36 patients were characterized by reproductive age (76.6%). From the collected data, 9 subjects (19.15%) were in menopause and 44 patients (93.62%) were sexually active. From the total of 47 subjects, 20 (42.55%) were tobacco consumers and 32 (68.09%) were physically active, in terms of practicing various sports.

**Table 1.** Demographic data and baseline characteristics of the study population.

| Baseline characteristics | Experimental |  |
| --- | --- | --- |
|  | n | % |
| **Age (years): mean [range]** | 47 | 41.23 [22 – 64] |
| **Gender** |  |  |
| Female | 47 | 100.00 |
| Male | 0 | 0.00 |
| **Ethnicity** |  |  |
| Caucasian | 47 | 100.00 |
| **Childbirth potential** |  |  |
| Yes | 36 | 76.60 |
| No | 11 | 23.40 |
| **Subject in menopause** |  |  |
| Yes | 9 | 19.15 |
| No | 38 | 80.85 |
| **History of cancer** |  |  |
| Yes | 2 | 4.26 |
| No | 45 | 95.74 |
| **Sexually active** |  |  |
| Yes | 44 | 93.62 |
| No | 3 | 6.38 |
| **Reproductive life** |  |  |
| Yes | 25 | 53.19 |
| No | 22 | 46.81 |
| **Tobacco consumption** |  |  |
| Yes | 20 | 42.55 |
| No | 27 | 57.45 |
| **Alcohol consumption** |  |  |
| Yes | 0 | 0.00 |
| No | 47 | 100.00 |
| **Physical activity** |  |  |
| Yes | 32 | 68.09 |
| No | 15 | 31.91 |

The vaginal smers prelevated at screening visit showed severely depressed *Lactobacillus* for 46.80% of the participants, numerous leukocytes for 78.72% of the participants and the increase in the parabasal epithelial cells for 68.08% of the participants. The cultures showed the dominant presence of *group B Streptococcus* and *Enterococcus* aerobes.

### 3.2. Change in vaginal symptoms

For 34 out of 47 participants (72.34%) treated with Cerviron^®^, the medical device had a beneficial effect on the score of vaginal symptoms, while for only 13 participants (27.66%) the score remained the same. Performing a test for equality of proportions with continuity correction, at 5% significance level, and considering a null hypothesis that is no statistically significant difference between baseline visit and final visit, we evidenced that the differences between these values were statistically significant (p < 0.001, 95% CI [57.42% - 87.25%]).

A very important aspect of Cerviron^®^ treatment is that it reduces the symptoms of vaginitis after 30 days. Thus, for 9 out of 47 patients (19.15%) who were treated, the medical device had a beneficial effect on vaginal symptoms score (p < 0.05, 95% CI [5.77% - 32.52%]) measured at visit 2 (30 days). This improvement in vaginal symptoms score is much more evident at 60 days when 25 out of 47 patients (53.19%) showed an improvement in symptoms (p < 0.001, 95% CI [36.80% - 69.58%]).

Even if between Visit 3 (60 days) and Visit 4 (90 days) there is a reduction of the score of 19.5%, more precisely the score improves for 9 patients, this improvement is not statistically significant (p = 0.081).

The treatment effect in terms of vaginal symptoms between visits is graphically presented in Figure 2.

**Figure 2.**
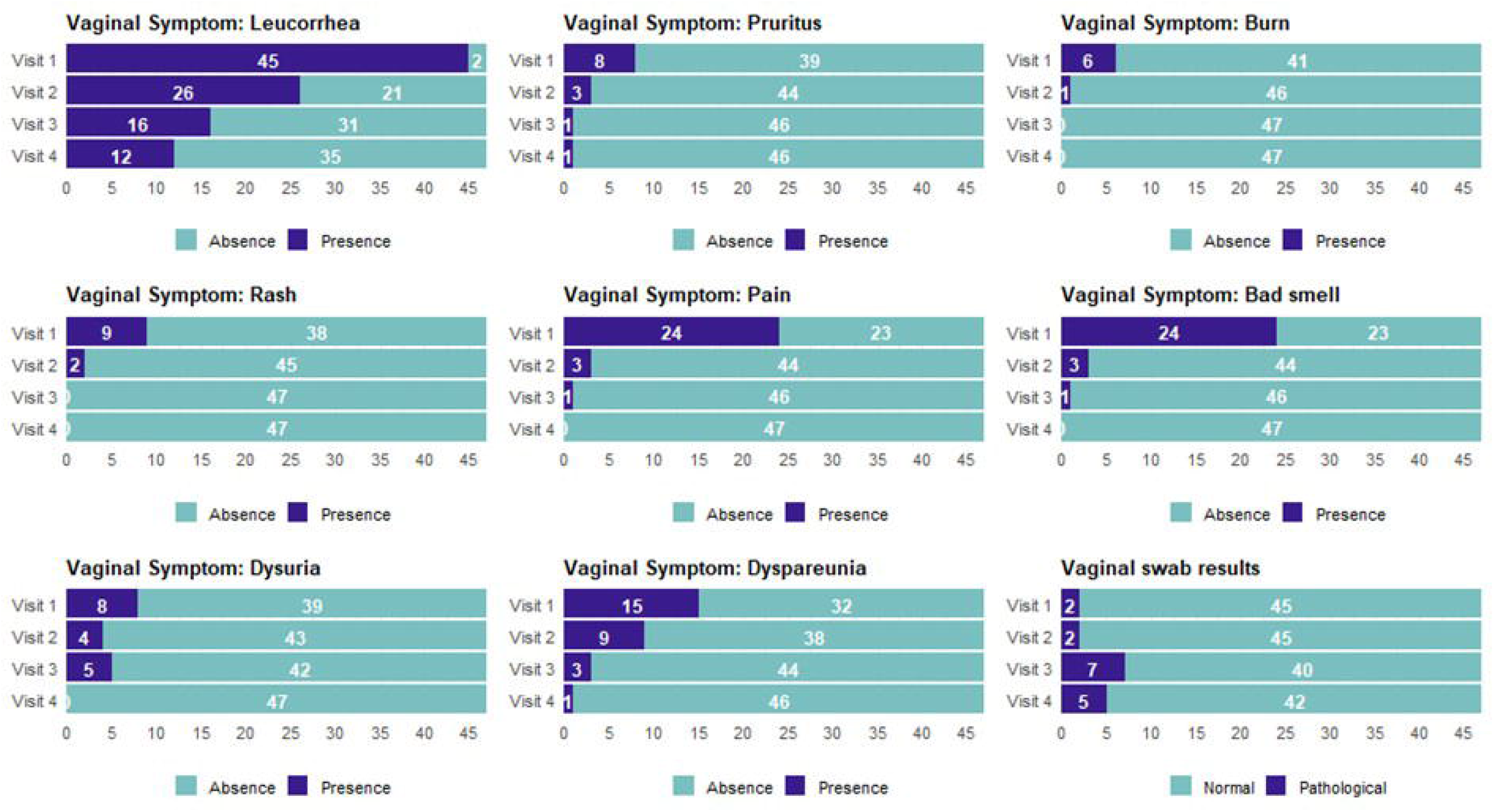
Change from baseline for vaginal symptoms - comparison between visits.

At the same time, treatment performance on each symptom was studied separately. Thus, it was observed that treatment for 90 days with Cerviron^®^ significantly reduced the following symptoms: leucorrhea (p <0.001), pruritus (p < 0.05), burn (p < 0.05), rash (p < 0.05), pain (p < 0.001), malodor (p < 0.05), dysuria (p < 0.05), and dyspareunia (p < 0.001).

The evolution of each symptom from baseline Visit, and throughout all the study visits is represented in Figure 3.

**Figure 3.**
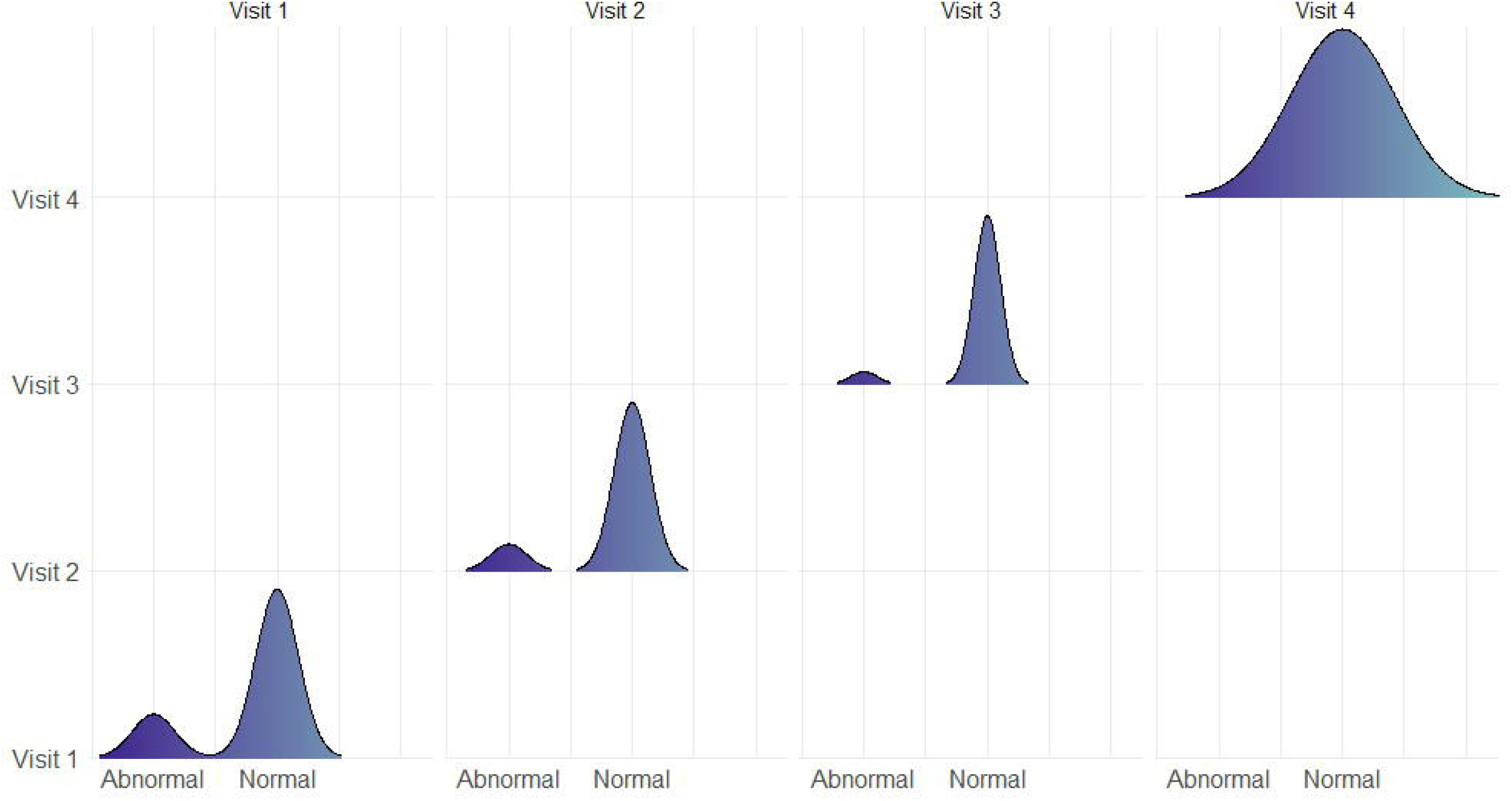
Vaginal symptoms and vaginal swab results - comparison between visits.

### 3.3 Change in vaginal discharge aspect

The difference in vaginal discharge aspect between baseline visit and vinal visit was statistically significant at 5% significance level (p < 0.05).

The vaginal discharge aspect is useful in the management of vaginal infections. The change in vaginal discharge aspect was assessed by the investigator as an objective finding and was verified by laboratory examination during every study visit. The vaginal discharge aspect improved during the medical device usage. While at the baseline visit, participants were evaluated with abundant discharge (N=4), moderate discharge (N=37) and mild discharge (N=6), after 90 days, some participants had no vaginal discharge (N=9), and other participants were reported with mild vaginal discharge (N=13) or moderate discharge (N=25). As such, Cerviron^®^ ovules proves its performance in improving the vaginal discharge aspect after 3 months of usage, as shown in Table 2.

**Table 2.** Change in vaginal discharge aspect score between each visit.

| Change in Vaginal Discharge Aspect | Visit 1 | Visit 2 | Visit 3 | Visit 4 |
| --- | --- | --- | --- | --- |
| Absent | 0 (0.00%) | 2 (4.26%) | 8 (17.02%) | 9 (19.15%) |
| Mild | 6 (12.77%) | 26 (55.32%) | 21 (44.68%) | 13 (27.66%) |
| Moderate | 37 (78.22%) | 19 (40.43%) | 18 (38.30%) | 25 (53.19%) |
| Abundant | 4 (8.51%) | 0 (0.00%) | 0 (0.00%) | 0 (0.00%) |
| Purulent | 0 (0.00%) | 0 (0.00%) | 0 (0.00%) | 0 (0.00%) |

### 3.4. Change in vaginal pH values

The vagina is essentially a dynamic microbial ecosystem with constantly adaptive vaginal pH levels. The vaginal pH level should be balanced between 3.8 – 4.5 (moderately acidic) in order to maintain a level of protection.

The vaginal pH can be affected by various factors such as age, antibiotics, vaginal hydration status, use of vaginal douching, unprotected sex, variations in the menstrual cycle, or nutrition [25]. In sexually active women, after unprotected intercourse, the vaginal defense system is hindered for 10-14 hours, due to the fact that the semen is alkaline [26].

During the present clinical investigation, a determination of pH values was performed at every 30 days. The difference in change of vaginal pH values between the baseline and final visit, at 5% significance level was statistically significant (p < 0.05).

The determination of pH values between study visits is showed in Table 3.

**Table 3.** Change in vaginal pH values between each visit.

| Change in vaginal pH values | Visit 1 | Visit 2 | Visit 3 | Visit 4 |
| --- | --- | --- | --- | --- |
| Normal | 38 (80.85%) | 41 (87.23%) | 44 (93.62%) | 47 (100.00%) |
| Abnormal | 9 (19.15%) | 6 (12.77%) | 3 (6.38%) | 0 (0.00%) |

It is observed that after 3 months of treatment with Cerviron^®^ all study participants returned to a normal, moderately acidic pH. These results indicate the effective role of the medical device in balancing the vaginal pH. Treatment performance was evaluated in the subset of sexually active participants. Figure 4 shows the evolution of pH in sexually active patients during the 90 days of treatment. After 30 days of treatment, a slight decrease was observed in cases with abnormal pH from 20.45% (N = 9) to 13.64% (N = 6). In contrast, at 60 days of treatment, the rate of patients with abnormal pH decreased to only 6.82% (N = 3), which shows that treatment with this medical device is effective (p < 0.05). At the end of the clinical investigation, all patients in this group had normal vaginal pH.

**Figure 4.**
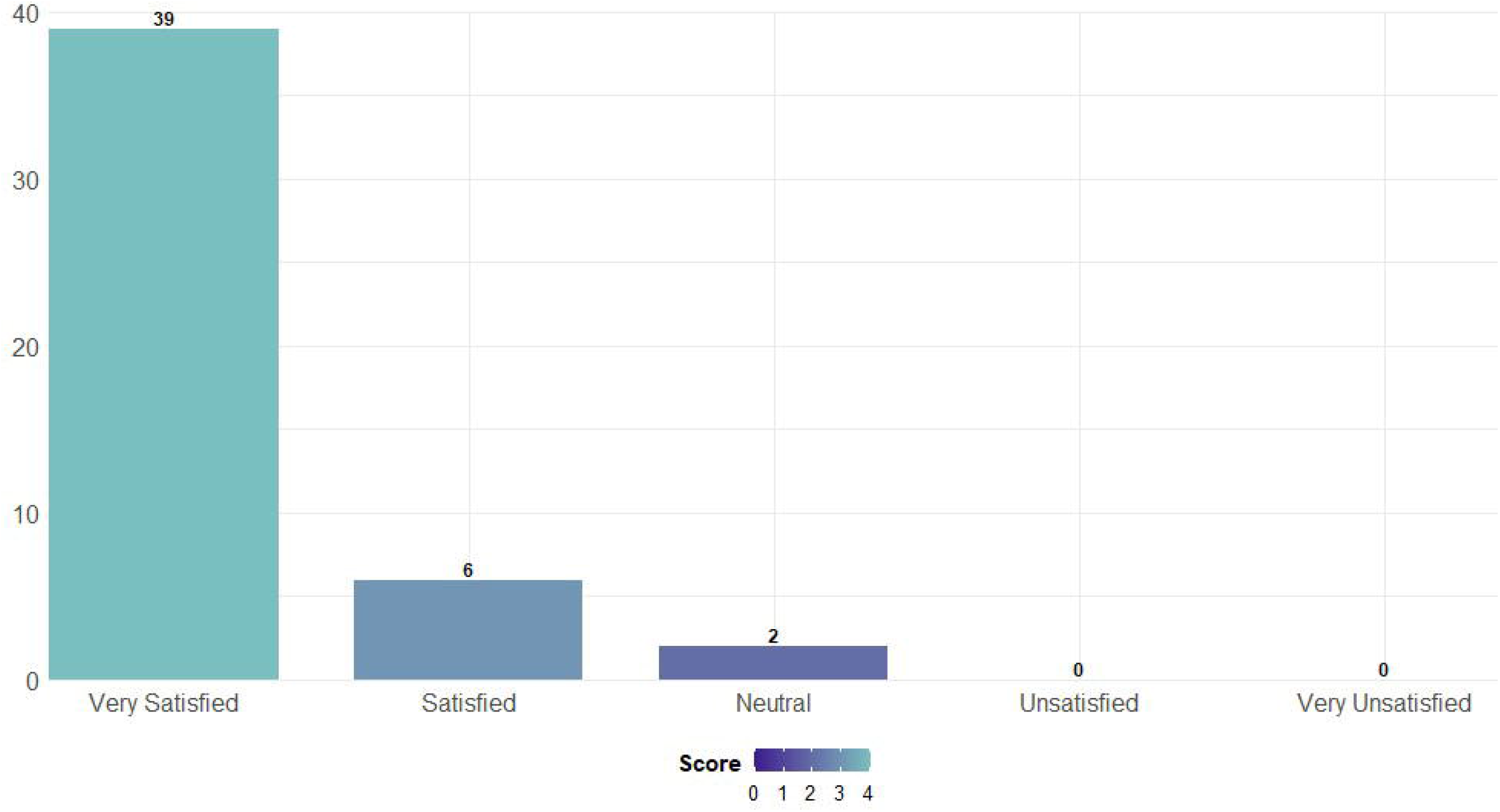
Changes in vaginal pH in sexually active study participants.

### 3.5. Change in vaginal inflammation

Within 3 months of treatment with the medical device, the changes in inflammatory and parabasal epithelial cells were relevant, with a number of 32 study participants recording an improvement.

McNemar’s Chi-squared test with continuity correction was performed to assess the difference of the vaginal inflammation score between the baseline and final visit (Visit 4). At a 5% significance level and considering a null hypothesis that is no statistically significant difference between baseline visit and final visit, we evidenced that the differences between these values were statistically significant (*p < 0*.*001*). The change in vaginal inflammation is represented in Table 4.

**Table 4.**
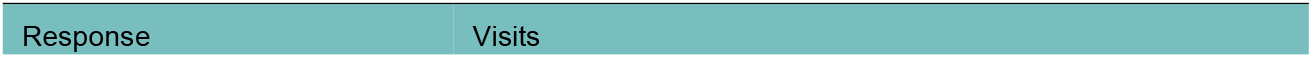
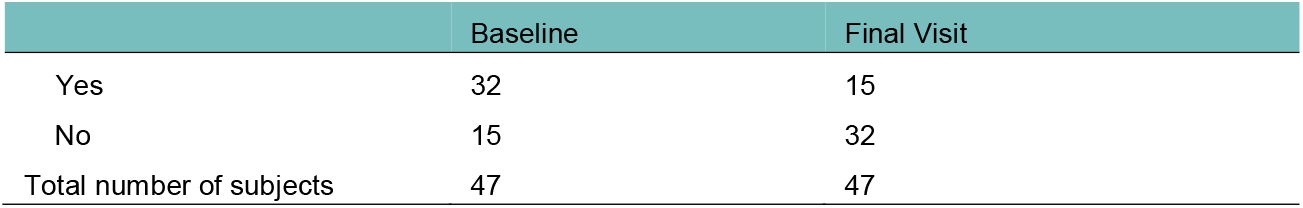
Change in vaginal inflammation score between baseline and final visit.

### 3.6. Degree of satisfaction after using Cerviron^®^

In terms of the degree of satisfaction offered, participants in the largest proportion (95.74%) said they were satisfied after using the medical device, as shown in Figure 5. Only 2 participants declared themselves neutral in terms of satisfaction.

**Figure 5.**
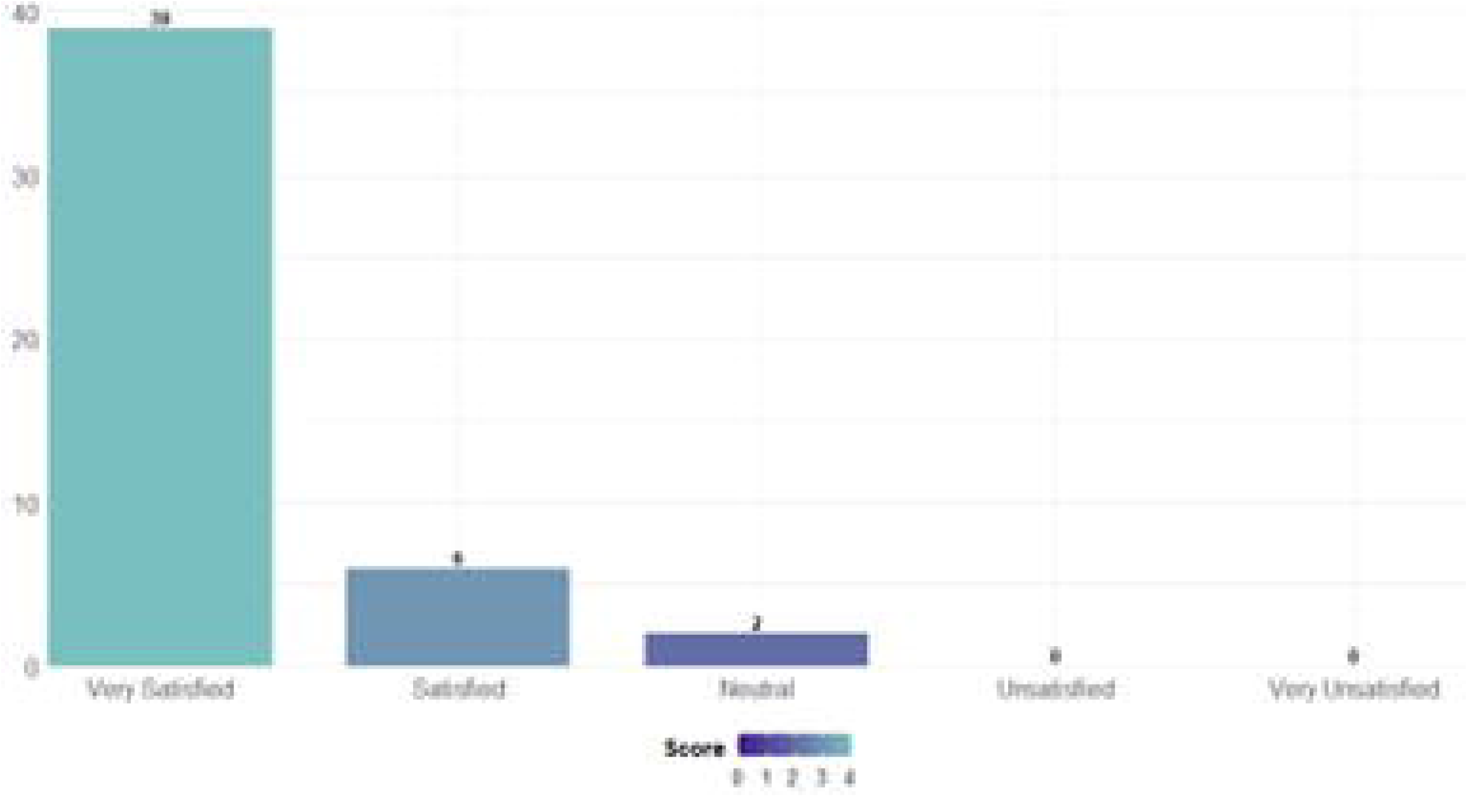
Participant Satisfaction Score – Likert Scale.

### 3.7. Safety of the medical device

During the study conduct, safety was assessed at every visit. Four study participants reported mild, grade-1 vaginal infections with *E. Coli* (N=2) and *group B Streptococcus* (N=2) during the study. Infection was resolved over time with oral antibiotics and the treatment with Cerviron^®^ continued, as per the treating physician recommendation.

At the end of study, no treatment-related adverse events were reported.

## 4. Discussion

A very important aspect in the management of non-specific vaginitis is maintaining the balance between endogenous vaginal bacteria. The vaginal fluid is maintained by *Lactobacilli* producing H_2_O_2_ and as such, preserving an acidic vaginal environment and a healthy vaginal ecosystem. The administration of broad-spectrum antibiotic use can trigger the alteration of the vaginal microflora leading to *Candida species* overpopulation and infection. Douching and unprotected vaginal intercourse can increase pH as well. Taking into account these premises, the medical device’s beneficial role in balancing the vaginal pH is very important. The medical device composition seems to provide multiple benefits, especially in the improvement of the symptoms associated with inflammation and infections. Bismuth subgallate is an insoluble product, with a very low bioavailability which creates a physical barrier over the affected area of the vaginal mucosa and therefore does not allow oxygen and pathogens to come in contact with it. In this way, it creates the premises that allow the damaged tissue to heal naturally. The role of exogenous collagen is to be a “ sacrificial substrate” in order to reduce excess matrix metalloproteinases that delay wound healing [27]. The phytotherapeutic extracts and hexylresorcinol are used as preservatives to prevent contamination with microorganisms.

Regarding sexual activity, Cerviron^®^ can be safely administered to both sexually active women as well as to abstinent women.

A recent study underlines the fact that the prevalence of aerobic vaginitis among pregnant women is four-times higher than in nonpregnant women [28]. As such, early diagnosis and treatment of aerobic vaginitis or its relapse before and during pregnancy are very important to reduce severe complications. The performance of the medical device in pregnant females will be a goal of future research.

Another potential recommendation could be represented by cervical and vaginal wounds, traumatic or secondary to surgical interventions, where hemorrhage, infection and inflammation represent serious clinical concerns. Bismuth subgallate has a local astringent action, forming a protective barrier in the vaginal mucosa which prevents the adhesion of pathogenic bacteria to the vaginal mucosa and promotes natural healing of damaged tissue. Histometric measurements in studies of bismuth subgallate-treated wounds showed a larger area of ulceration of bismuth subgallate-treated wounds on the first day, which could be explained by the presence of bismuth subgallate that fills the wound to the tip and acts as a barrier to the initial contraction. This filling could be a factor that helps protect the wounds from trauma and bacteria [29][30]. The oral and vaginal mucosa are very similar in terms of structure and permeability, both composed of stratified squamous epithelium that is not keratinized, and with similar stromal characteristics, therefore we would expect a similar response.

The adverse effects should not occur during Cerviron^®^ administration. Bismuth salts used for indications like gastrointestinal disorders or syphilis have been associated with some adverse effects, especially dermatologic reactions, but used in much higher dosages and exposure [31]. The exposure to topical bismuth is minimal in the composition of the medical device.

Our study has particular limitations that include the absence of a control group due to the unavailability of similar medical devices and a limited timeframe of only 3 months. Larger studies to detect the recurrences in an extended timeframe and to include a more diverse population should be designed to validate the conclusions of our study. Also, the exceedingly high rate of tobacco users should be the subject of future correlations between cigarette smoking and susceptibility to urogenital infections. Cigarette smoking has been associated with both the diagnosis of bacterial vaginosis and a vaginal microbiota lacking protective *Lactobacillus* but the mechanism linking smoking with vaginal microbiota is still unclear [32].

## 5. Conclusions

Cerviron^®^’s clinical performance as a stand-alone treatment in symptomatic, non-specific vaginitis was demonstrated throughout the present clinical investigation. Cerviron^®^ significantly alleviates the symptoms in aerobic vaginitis while restoring the normal pH and reducing signs of vaginal inflammation. Applied once a day for a sequence of 15 consecutive days, during 3 consecutive months, Cerviron^®^ may be a suitable treatment option in cases of aerobic vaginitis, in both women of reproductive age and postmenopausal women. At the final study visit, no study participants presented treatment-related adverse events, which makes the medical device safe to administer. The medical device is considered a safe and effective alternative to alleviate specific signs and symptoms of atrophic vaginitis in postmenopausal women (dysuria, dyspareunia and vaginal inflammation), especially when hormonotherapy is not recommended. Some limitations of this study include the absence of a control group, a short treatment follow-up, the heterogenicity of the sample population and also the exceedingly high rate of tobacco users.

It is interesting to note that Cerviron^®^ reduces the symptoms of vaginitis after 30 days, showing beneficial effect on the score of vaginal symptoms. It must also be considered that, since there were no adverse events related to the usage of the medical device, the treatment might be applied whenever deemed necessary.

Its clinical efficacy and safety in microlesion repair and postoperative care in patients undergoing local excision of cervical lesions in currently under evaluation (protocol code CYRON/02/2021, NCT04735718). Future research is needed to confirm its antibiotic boosting effect in patients with mixed aerobic infections requiring anti-infectious treatment regimens and its tolerability profile during pregnancy.

## Data Availability

All data produced in the present study are available upon reasonable request to the authors.

## Acknowledgments

Perfect Care Distribution provided financial support for the clinical investigation. However, Perfect Care Distribution had no involvement in the study design, the collection, analysis and interpretation of data, in the writing of the report and in the decision to submit the article for publication.

The authors thank the study participants that voluntarily enrolled in the study. The authors also thank Alexandru-Remus Pinta for his work provided during the statistical analysis and Adrian Pocola for technical support provided during data management collection. All authors approved the final version of the manuscript to be published.

## Data sharing

Perfect Care Distribution provides access to all individual participant data collected during the trial, after anonymization. Access is provided after a proposal has been approved by an independent review committee identified for this purpose and after receipt of a signed data-sharing agreement. Data and documents, including the clinical investigation plan, clinical study report, and blank or annotated case report forms, will be provided in a secure data-sharing environment.

